# Performance characteristics of highly automated HSV-1 and HSV-2 IgG testing

**DOI:** 10.1101/2024.02.15.24302848

**Authors:** Katharine H. D. Crawford, Stacy Selke, Gregory Pepper, Erin Goecker, Aniela Sobel, Anna Wald, Christine Johnston, Alexander L. Greninger

## Abstract

Herpes simplex virus (HSV) infections are one of the most common and stigmatized infections of humankind, affecting more than 4 billion people around the world and more than 100 million Americans. Yet most people do not know their infection status and antibody testing is not recommended, partly due to poor test performance. Here, we compared the test performance of the Roche Elecsys HSV-1 IgG and HSV-2 IgG, DiaSorin LIAISON HSV-1/2 IgG, and Bio-Rad BioPlex 2200 HSV-1 & HSV-2 IgG assays with the gold-standard HSV western blot in 1994 persons, including 1017 persons with PCR or culture-confirmed HSV-1 and/or HSV-2 infection. Across all samples, the Bio-Rad and Roche assays had similar performance metrics with low sensitivity (<85%), but high specificity (>97%) for detecting HSV-1 IgG and both high sensitivity (>97%) and high specificity (>98%) for detecting HSV-2 IgG. The DiaSorin assay had a higher sensitivity (92.1%) but much lower specificity (88.7%) for detecting HSV-1 IgG and comparatively poor sensitivity (94.5%) and specificity (94.2%) for detecting HSV-2 IgG. The DiaSorin assay performed poorly at low-positive index values with 60.9% of DiaSorin HSV-1 results and 20.8% of DiaSorin HSV-2 results with positive index values <3.0 yielding false positive results. Based on an estimated HSV-2 seroprevalence of 12% in the United States, positive predictive values for HSV-2 IgG were 96.1% for Roche, 87.4% for Bio-Rad, and 69.0% for DiaSorin, meaning nearly 1 of every 3 positive DiaSorin HSV-2 IgG results would be falsely positive. Further development in HSV antibody diagnostics is needed to provide appropriate patient care.

## INTRODUCTION

Herpes simplex virus types 1 and 2 are important human pathogens that cause lifelong infections characterized by latent infection with cycles of viral mucosal replication. Classically, HSV infections present with episodes of recurrent, painful lesions of the oral or genital epithelium, but many infected persons remain asymptomatic.^1^ HSV is characterized by two types: HSV-1 is near-ubiquitous and infects an estimated 67% of the world’s population and ∼48% of adults in the United States of America.^2,3^ HSV-1 is associated with oral lesions, but is an increasingly recognized cause of genital herpes.^4^ Conversely, HSV-2 is classically associated with genital herpes and infects approximately 13% of adults worldwide and ∼12% of adults in the United States of America.^2,3^

HSV infections can be diagnosed by detecting the virus itself—typically through culture or PCR—or detecting antibodies against viral proteins.^5^ The HSV lifecycle, which includes cycles of lytic and latent infection, complicates diagnosis and limits accurate PCR testing to episodes of active mucosal infection. Not all episodes of active mucosal infection are symptomatic and high rates of asymptomatic shedding mean symptoms alone cannot be used to guide testing to prevent transmission.^6^ High-throughput, sensitive and specific serological diagnostics are required for both clinical care and for potential screening to diagnose the vast majority of HSV infections that currently go undetected.^7^

Most high-throughput HSV serology assays detect type-specific IgG responses to the HSV-1 or HSV-2 glycoprotein G (gG_1_ for HSV-1 and gG_2_ for HSV-2), which share only ∼40% amino acid identity.^8^ However, IgG to gG_1_ and gG_2_ may still be cross-reactive and insufficiently specific, leading the United States Preventive Task Force to recommend against screening for genital herpes infections among asymptomatic adolescents and adults.^9^ CDC Sexually Transmitted Infection (STI) Treatment Guidelines recommend a two-step testing process, using a sensitive enzyme/chemiluminescence assay for screening followed by a second, more specific confirmatory assay.^10^ The gold standard for confirmatory serological diagnosis of HSV-1 and HSV-2 infections is a western blot that detects serological responses to multiple proteins within a viral lysate.^11^ However, this assay is laborious, and thus costly and only available at one reference lab. Here, we examined the sensitivity and specificity of three fully automated, high-throughput, type-specific HSV-1/2 IgG assays compared to the HSV western blot in two clinical cohorts comprising 1994 individuals.

## METHODS

### Cohorts

#### HSV Western Blot Serum Specimen Remnant Cohort

The initial cohort consisted of samples selected from the University of Washington (UW) HSV western blot (WB) sample remnants collected from January 2022 through April 2023. Sample selection was restricted to the 5423 samples collected within the UW Medicine system to ensure availability of clinical metadata, to approximate a more typical HSV testing population (since all UW specimens are tested only by WB without a prior screening test), and to avoid biasing specimens toward indeterminate results sent to our reference lab for WB confirmatory testing. Based on the WB results, we randomly selected 449 samples that were negative for both HSV-1 and HSV-2 antibodies, 177 samples that were positive for HSV-1 and negative for HSV-2 antibodies, 182 samples that were positive for HSV-2 and negative for HSV-1 antibodies, and 184 samples that were positive for both HSV-1 and HSV-2 antibodies. For each WB category, each sample came from a different person. Two persons contributed two samples as their WB serologic status changed during the sample selection timeframe and their samples were randomly selected in two result categories. Due to low sample volume, 13 of the originally selected samples were unable to be tested leaving 979 samples from 977 persons. Additionally, due to low sample volume and rare barcode errors, not all samples were able to be run on all instruments. The number of samples from this cohort run on each instrument and their concordance with the WB result are presented in Table 1. Use of excess clinical testing specimens was approved by the UW Institutional Review Board with a consent waiver (STUDY00010205).

**Table 1:** Number of samples from the clinical testing remnant sample cohort that were tested on each automated instrument per western blot (WB) category. Some samples were unable to be run on all instruments due to different volume requirements or rare barcode errors. The percent concordance with the WB results for each assay is also shown. For the Roche HSV-1 and HSV-2 assays, sample numbers are provided for each assay (e.g. HSV-1 sample number, HSV-2 sample number) since some samples had insufficient volume to be run on both assays.

| Western Blot Category | Number Tested: All assays | Number Tested: Bio-Rad BioPlex 2200 HSV-1 & HSV-2 IgG | % Concordance: Bio-Rad |  | Number Tested: DiaSorin LIAISON HSV-1 & HSV-2 IgG | % Concordance: DiaSorin |  | Number Tested: Roche Elecsys HSV-1 IgG, HSV-2 IgG | % Concordance: Roche |  |
| --- | --- | --- | --- | --- | --- | --- | --- | --- | --- | --- |
|  |  |  | HSV-1 | HSV-2 |  | HSV-1 | HSV-2 |  | HSV-1 | HSV-2 |
| HSV-1 neg & HSV-2 neg | 444 | 434 | 99.3% | 99.5% | 443 | 91.4% | 94.8% | 435, 427 | 100% | 100% |
| HSV-1 pos, HSV-2 neg | 175 | 172 | 88.4% | 96.5% | 175 | 93.1% | 92.0% | 173, 171 | 86.1% | 98.8% |
| HSV-1 neg, HSV-2 pos | 182 | 181 | 95.6% | 93.4% | 182 | 82.4% | 91.2% | 169, 165 | 95.3% | 97.0% |
| HSV-1 pos & HSV-2 pos | 178 | 178 | 85.4% | 88.8% | 178 | 94.4% | 78.1% | 168, 168 | 85.7% | 95.2% |
| Total | 979 | 965 | 94.1% | 95.9% | 978 | 90.6% | 90.6% | 945, 931 | 94.1% | 98.4% |

#### PCR- or Culture-confirmed Cohort

A second cohort consisted of serum specimens from persons with genital lesions PCR- or culture-positive for HSV-1 or HSV-2 followed by the UW Virology Research Clinic who provided written informed consent approved by the UW IRB.^12,13,14^ We selected one serum specimen each from 1074 HIV-negative individuals who tested positive for HSV-1 or HSV-2 by PCR and/or culture and were confirmed seropositive for the corresponding HSV type by WB. Some persons with HSV-2 positive genital lesions had both HSV-2 and HSV-1 antibodies when tested with the WB, resulting in 131 samples positive for HSV-1 only with both viral and WB testing, 566 samples positive for HSV-2 only with both viral and WB testing, and 320 samples positive for HSV-2 with viral testing and both HSV-1 and HSV-2 antibodies on WB. Virological data on shedding rates in genital herpes infections (measured as percent of swabs positive for HSV-1 or HSV-2 by non-type-specific PCR over total number of swabs) were also available from 315 of 1017 persons in the VRC cohort (Supplemental Data).^15^

### Assay Comparison

The DiaSorin Liaison HSV-1 and HSV-2 IgG chemiluminescent immunoassays (CLIA) (K081685 & K081687), the Roche cobas Elecsys HSV-1 and HSV-2 IgG sandwich electrochemiluminescence immunoassays (ECLIA) (K120625 & K121895), and the Bio-Rad BioPlex HSV-1/2 IgG multiplexed bead immunoassay (K120959) were compared to the HSV-1 and HSV-2 WB performed at the UW Virology lab. These commercially manufactured assays are widely used fully automated HSV serology assays that are FDA 510(k) cleared for clinical use in the United States. QC and calibration were performed for each assay according to laboratory procedures for HSV WB or manufacturer’s instructions for commercial assays.

#### Western Blot

All samples were previously tested for HSV-1 and HSV-2 antibodies by WB at UW as part of clinical testing or research.^11^ In this assay, HSV-1 and HSV-2 proteins from lysates of HSV-infected cells are separated by gel electrophoresis then transferred to nitrocellulose paper to make HSV type-specific blots containing separated, fixed proteins from either HSV-1 or HSV-2. These blots are then incubated with a 1:50 dilution of the patient’s serum and antibodies that bind the viral proteins are detected by an enzyme-mediated color change. The staining pattern of the antibodies in the patient’s serum on each blot allow for type-specific determination of HSV antibodies and determination of a patient’s serologic status. Blots are read by three independent readers and reader consensus is used to call samples positive, negative, or indeterminate for HSV-1 and HSV-2 antibodies. Notably, if any reader finds the initial blot indeterminate, samples are adsorbed for HSV-1 and HSV-2 antibodies and re-run or repeated with more sample volume.^11^ Following adsorbing or repeating the sample, results are re-read by three independent readers and consensus-called as positive, negative, or indeterminate. No samples with a final WB result of indeterminate were included in this study. Following initial clinical testing, samples were stored at −10 to −25°C until use in this study (average of 12 months).

#### DiaSorin LIAISON HSV-1 and HSV-2 Type Specific IgG Assays

Samples were thawed and briefly mixed prior to testing with the DiaSorin LIAISON HSV-1 and HSV-2 IgG CLIA according to manufacturer instructions (DiaSorin, United States). In this assay, magnetic beads coated with recombinant antigens specific for either HSV-1 glycoprotein (gG1) or HSV-2 glycoprotein (gG2) are incubated with patient serum. After washing, the beads are incubated with an isoluminol-conjugated anti-human IgG mouse monoclonal antibody. Another wash removes any unbound antibody prior to inducing a flash chemiluminescence reaction. The resulting light signal, measured as relative light units (RLUs), indicates how much isoluminol-antibody conjugate and, thus, HSV-1 or HSV-2 IgG is present in the sample. The RLUs are converted to an index value using the on-board calculation based on the manufacturer-provided calibrators. This index value is used to categorize samples as negative (index *<*0.9), equivocal (index 0.90-1.09), or positive (index ≥1.1). No reportable range is listed on the package insert. The reported index values in this study ranged from <0.01 to >62.2 for the DiaSorin HSV-1 assay and from 0.03 to >23.6 for the HSV-2 assay. Linearity was confirmed for index values between 0.1-47 for HSV-1 and 0.15-21 for HSV-2 (Figure S1, Supplemental Data 2). Samples that were initially reported as equivocal were repeated at least twice and the median measurement was reported as the final result. For the VRC cohort, the equivocal samples were repeated at the end of the study, following an additional freeze/thaw. Following testing on the DiaSorin instrument, samples were stored at 4°C for *<*48 hours prior to testing using the Roche Elecsys assays.

#### Roche Elecsys HSV-1 IgG and HSV-2 IgG Assays

All samples were tested using the Roche Elecsys HSV-1 and HSV-2 IgG immunoassays according to manufacturer instructions. In this assay, serum samples are incubated with biotinylated recombinant HSV-1 gG1 or HSV-2 gG2 antigens and the same antigens labeled with a ruthenium complex to form a double-antigen sandwich complex. Streptavidin-coated magnetic microparticles are then added and the sandwich complex binds to this solid phase through the interaction of biotin and streptavidin. The microparticles are then magnetically captured on the surface of an electrode and all unbound substances are washed away. Applying a voltage to the electrode induces chemiluminescence of the ruthenium complex which is measured by a photomultiplier. On-board software converts this electrochemiluminescent signal to a cutoff index (COI) based on manufacturer-supplied calibrators. A COI *<*1.0 is considered non-reactive (negative) and a COI ≥1.0 is reactive (positive), with no equivocal range. Neither low nor high values are censored as > or < and reported values ranged from a COI of 0.026 to 234.8 for HSV-1 and 0.083 to 575.1 for HSV-2. Linearity was confirmed from 0.1-140 for HSV-1 and 0.2-350 for HSV-2 (Figure S1, Supplemental Data 2). After testing using the Roche Elecsys assays, samples were frozen at −20°C for 2-3 weeks prior to running on the Bio-Rad BioPlex instrument.

#### Bio-Rad BioPlex 2200 HSV-1 and HSV-2 IgG Assay

Serum samples were thawed, mixed, and briefly spun (700xg for 4 min) prior to testing with the Bio-Rad BioPlex 2200 HSV-1 and HSV-2 IgG assay according to manufacturer instructions. The BioPlex assay is a multiplexed microparticle immunoassay. Briefly, serum samples are incubated with dyed beads coated with either recombinant HSV-1 gG1 antigen or a synthetic HSV-2 gG2 peptide. After washing, anti-human IgG antibody conjugated to phycoerythrin is added and the specimens are incubated at 37°C. After another wash cycle, the mixture passes through a detector that measures fluorescence of both the bead dye and phycoerythrin. This allows for simultaneous determination of antibody amount and antigen type (e.g. HSV-1 or HSV-2). Relative fluorescence intensity values are converted to antibody index (AI) using the on-board software and manufacturer-provided calibrators. An AI of *<*0.9 is negative, 0.9-1.0 is equivocal and ≥1.1 is positive. Results with indices less than 0.2 or greater than 8.0 are reported as <0.2 or >8.0, respectively. With this narrow measurement range, only 31% of results in this study had a non-bounded numerical result (i.e. from 0.2-8.0). Linearity was confirmed across the measurement range (0.2 to 8.0 AI) (Figure S1, Supplemental Data 2). Bio-Rad recommends obtaining an additional sample and repeating testing for patients with equivocal results. We could not test additional samples but did repeat all equivocal specimens at least twice to assess the reproducibility of these equivocal results. Of the 35 initially equivocal samples, 25 were repeated after an additional freeze-thaw cycle. For repeated samples, the median result was accepted as the final result.

#### Data analysis and availability

Data were analyzed using R-Studio.^16^ Results that remained equivocal after repeating were not included in calculations of sensitivity and specificity. Line-item testing data are provided in the supplemental data with coded sample identifiers.

## RESULTS

### Clinical Testing Remnant Samples Cohort

#### Samples and Demographics

HSV-1 and HSV-2 IgG were measured in 979 remnant Western blot samples from 977 individuals from all four WB result categories: 444 negative for both HSV-1 and HSV-2 IgG, 175 positive for HSV-1 but negative for HSV-2 IgG, 182 positive for HSV-2 IgG but negative for HSV-1 IgG, and 178 positive for IgG to both HSV-1 and HSV-2. For each instrument, we calculated concordance with the WB results (Table 1).

The clinical remnant samples consisted of 474 (54%) women and 503 (46%) men with a median age of 43 years (range: 3-91 years, Table 2). The age distribution was bimodal, which was likely related to the two separate major indications for HSV serology testing—STI testing and pre-transplant screening (Figure S2, Table 2). Most people identified as white and non-Hispanic (64.3%) and were from Washington state (87.1%).

**Table 2:** Demographics for the clinical testing sample remnant cohort.

|  | Clinical Testing Cohort<br>n=977 |
| --- | --- |
| **Median age in years (range)**<br>≤ 45 years old (%)<br>> 45 years old (%) | **43 (3-91)**<br>527 (53.9%)<br>450 (46.1%) |
| **Sex in medical record**<br>Female (%)<br>Male (%) | 474 (48.5%)<br>503 (51.5%) |
| **Race and ethnicity**<br>Hispanic, All Races<br>Non-Hispanic, Asian<br>Non-Hispanic, Black or African American<br>Non-Hispanic, White | 81 (8.3%)<br>108 (11.1%)<br>75 (7.7%)<br>628 (64.3%) |
| **State**<br>Washington<br>Alaska, Idaho, or Oregon<br>Other | 851 (87.1%)<br>65 (6.7%)<br>61 (6.2%) |
| **Top 5 order diagnosis groups**<br>STI screening<br>End stage renal disease<br>Organ donor<br>Lymphoma<br>Multiple myeloma | 251 (25.7%)<br>106 (10.8%)<br>59 (6.0%)<br>51 (5.2%)<br>45 (4.6%) |

#### Comparison to WB for Clinical Testing Sample Remnants

No single assay performed best for both HSV-1 and HSV-2 in the clinical remnant cohort. The DiaSorin HSV-1 IgG assay had the highest sensitivity (94.8%), but lowest specificity (90.4%), while both the Roche and Bio-Rad assays had lower sensitivities (85.9% and 87.1%, respectively), but higher specificities (98.7% and 98.2%, respectively, Table 3). For measuring HSV-2 IgG, the Roche assay had the highest sensitivity (96.1%) and specificity (99.7%). The Bio-Rad HSV-2 IgG assay had a sensitivity of 91.9% and specificity of 98.8%, and the DiaSorin HSV-2 IgG assay was both the least sensitive (85.4%) and least specific (94.5%).

**Table 3:**
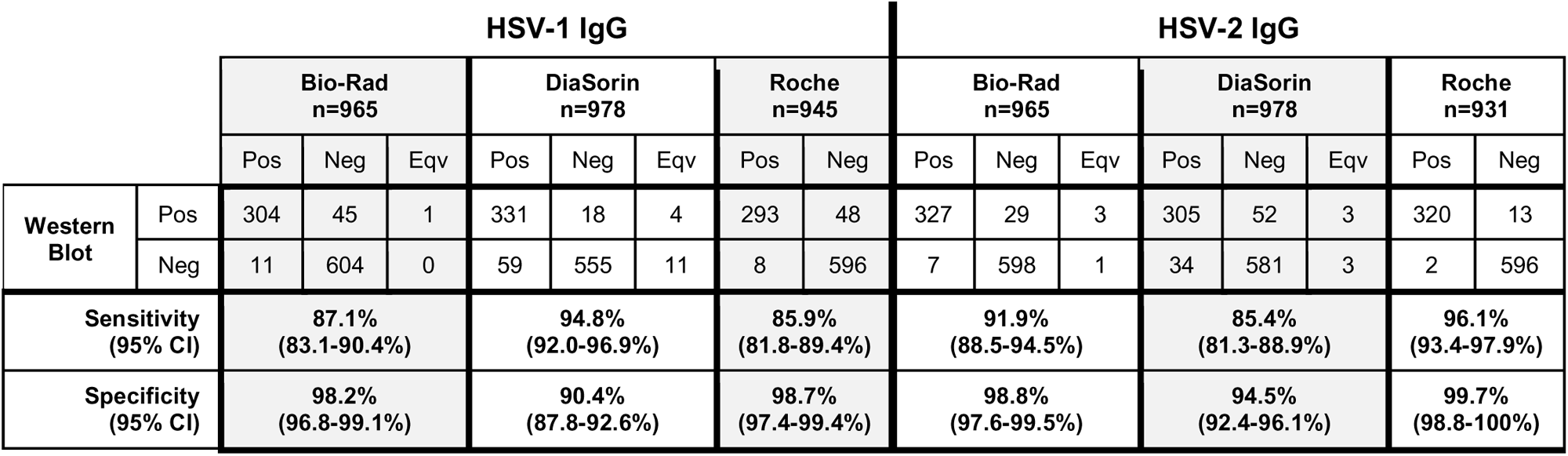
Test results for each automated instrument compared to the western blot results for the clinical testing remnant sample cohort. Equivocal (Eqv) results were not included in calculating sensitivity and specificity.

On initial testing, 19 results (1.0%; 8 HSV-1, 11 HSV-2) were equivocal on the Bio-Rad assay and 24 results (1.2%; 16 HSV-1, 8 HSV-2) were equivocal on the DiaSorin assay. For the Bio-Rad assay, the majority of equivocal results—88% for HSV-1 and 64% for HSV-2—were revised to a non-equivocal final result of which 57% of the revised results were concordant with WB. Only 12.5% of the DiaSorin equivocal results had a non-equivocal final result, but all revised results were concordant with WB (see Supplemental Material for detailed discussion). Only two samples were equivocal on more than one assay. One sample was HSV-1 IgG positive only by WB and had initially equivocal results on the Bio-Rad HSV-1 IgG assay and the DiaSorin HSV-2 IgG assay. After repeating, this sample was correctly positive for HSV-1 IgG on the Bio-Rad instrument, but remained equivocal for HSV-2 IgG on the DiaSorin instrument. The other sample was positive for both HSV-1 and HSV-2 IgG by WB, but equivocal for both analytes on the DiaSorin instrument and remained equivocal after repeating. Overall, equivocal results were more platform-specific than sample-specific as there were no samples that were equivocal for the same analyte on multiple instruments.

#### Investigation of Discordant Results

We next examined whether the same specimens had discordant results compared to WB on multiple automated instruments. In total, 201 samples (20.5% of samples) had results discordant with the WB result on at least one instrument for at least one analyte (e.g. HSV-1 IgG or HSV-2 IgG). Of these 201 samples, 102 (51%) were discordant for HSV-1 only, 78 (39%) were discordant for HSV-2 only, and 21 (10%) were discordant for both assays on at least one instrument. Since 21 samples were discordant with the WB results for both HSV-1 and HSV-2, in total 222 assay results were discordant with the WB results on at least one automated instrument. Notably, 13 of these results came from samples that were not run on all 3 instruments (Table 1). Excluding those, 134 results were discordant on one instrument only, 47 were discordant on 2 tests, and 28 were discordant with the WB results on all 3 tests (Figure 1).

**Figure 1:**
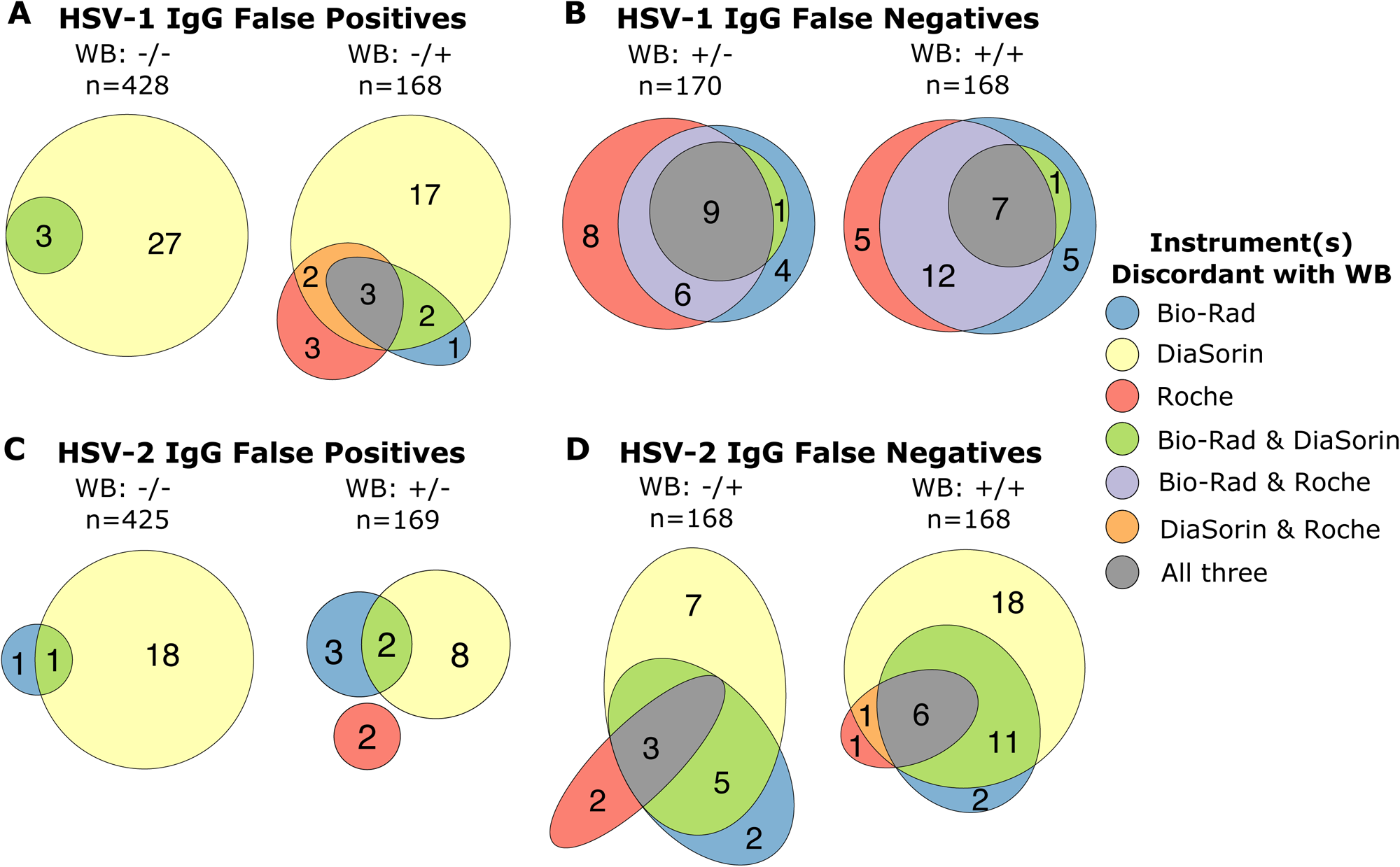
Euler plots showing the number of samples that disagree with the western blot results for each automated instrument colored by which instruments were discordant with the WB results. Data are presented separately for each WB category with the HSV-1 WB result left of the “/” and the HSV-2 WB result right of the slash. Only samples that were run on all three instruments are included with n showing the number of samples tested on all three instruments for the given WB category. Within each category, the size of each colored section is roughly proportional to the number of samples contained within it. A) Number of samples that were negative for HSV-1 IgG by WB, but falsely positive on the automated instruments. B) Number of samples that were positive for HSV-1 IgG by WB, but falsely negative on the automated instruments. C) Number of samples that were negative for HSV-2 IgG by WB, but falsely positive on the automated instruments. D) Number of samples that were positive for HSV-2 IgG by WB, but falsely negative on the automated instruments. Plots made using the eulerr R package.^22^

We found the false positivity rate for HSV-1 or HSV-2 antibodies was higher in samples that were positive for antibodies to the other virus. For HSV-1, 17% (31 of 182) of samples that were seropositive for HSV-2 only by WB tested falsely positive for HSV-1 IgG on at least one automated instrument compared with 7.4% (33 of 444) of samples that were negative for any HSV antibodies by WB (two sample z-test: p-value <0.001). There was a similar trend for HSV-2 as 9.7% (17 of 175) of samples that were positive for HSV-1 only by WB tested falsely positive for HSV-2 IgG on at least one automated instrument, while only 5.2% (23 of 443) of samples that were negative for all HSV antibodies by WB tested falsely positive for HSV-2 IgG (two sample z-test: p-value=0.06). Notably, 76% of HSV-1 false positive and 74% of HSV-2 false positive test results came from the the DiaSorin instrument. For HSV-1, the DiaSorin instrument had fewer false negative results than the other instruments and there were no samples where it was the only instrument to falsely call a sample negative (Figure 1). However, for HSV-2, the DiaSorin instrument had numerous false negative results and 43% (25 of 58) of the HSV-2 false negative results were unique to the DiaSorin instrument.

During WB testing, 168 of the 979 (17.2%) samples were repeated with a larger sample volume or adsorbed for HSV-1 and HSV-2 antibodies and re-tested. These samples that required repeating or absorption were significantly more likely to give discordant results on the automated instruments (two-sample z test: p-value <0.001). Of the 168 samples that were absorbed or repeated, 46% disagreed with the WB results on at least one automated instrument. In contrast, among the 811 samples that were not adsorbed or repeated only 15% of samples were discordant.

#### Quantitative Results

Although each assay’s primary result is positive, negative, or equivocal (for the Bio-Rad and DiaSorin assays) for HSV-1 or HSV-2 IgG, each automated assay also provides a quantitative index value. Previous studies have found that “low positive” index values of <3.0 (Figure 2, red line) are associated with more false positive results and a decreased positive predictive value.^14^ Indeed, when samples are grouped by index value, samples with positive index values <3.0 have the highest percent of false positive results for both HSV-1 and HSV-2 IgG on all instruments (Figure 3). This is especially true for the DiaSorin instrument where 76.1% of low-positive index values are falsely positive for HSV-1 IgG and 38.7% of low-positive results are falsely positive for HSV-2 IgG.

**Figure 2:**
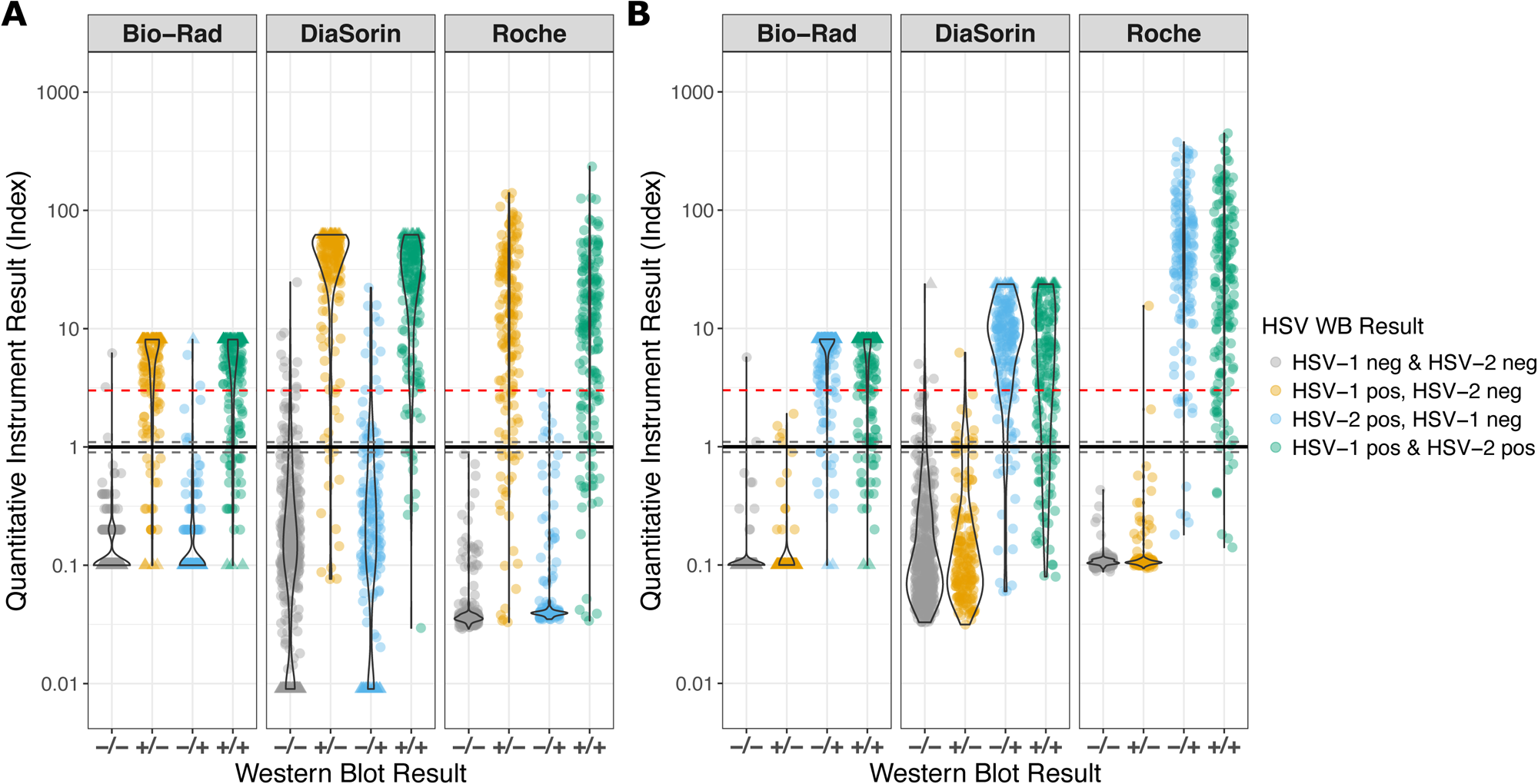
Comparison of quantitative assay results for each instrument and known western blot category for the clinical testing sample remnant cohort. A) HSV-1 IgG test results. B) HSV-2 IgG test results. The black line shows the cutoff value of 1.0 with the grey dotted lines at 0.9 and 1.1 delineating the index values that are called as equivocal by the Bio-Rad and DiaSorin instruments. The red dotted line at 3.0 marks a common cutoff for “low-positive” results. Values outside the reportable range for the Bio-Rad and DiaSorin instruments were assigned values 0.1 above or below the cutoff value and are shown as triangles.

**Figure 3:**
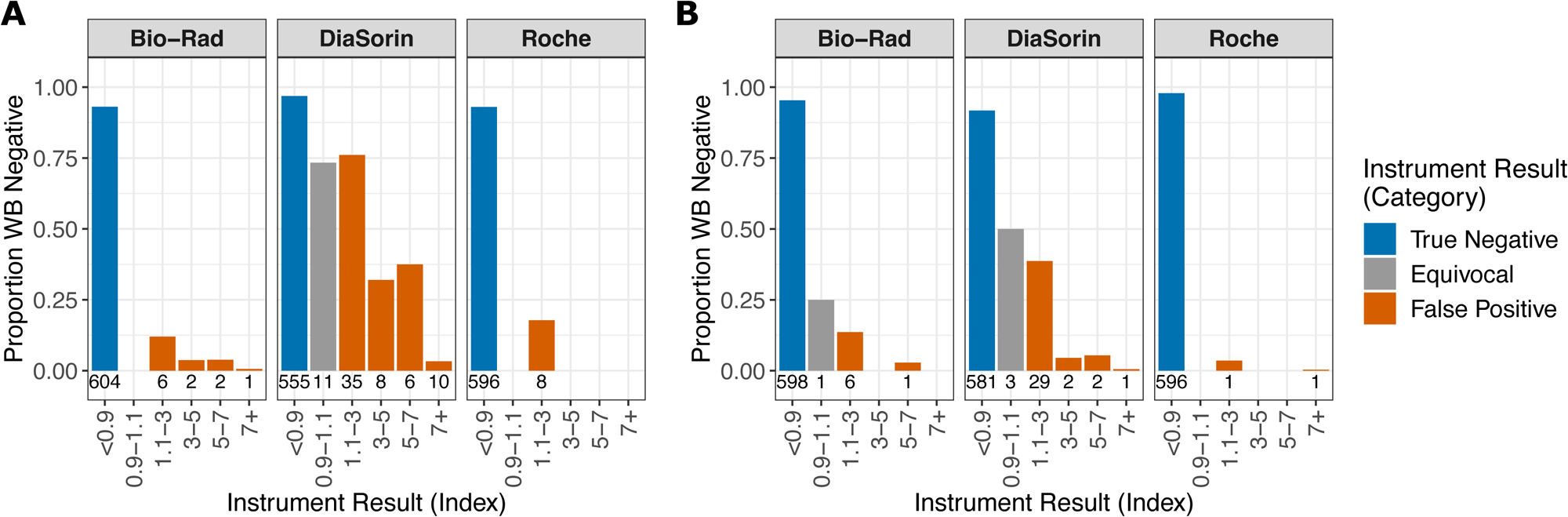
Percent of results that are negative by western blot for each index value bin for each instrument for the clinical testing remnant sample cohort. A) HSV-1 IgG. B) HSV-2 IgG. All samples to the right of 1.1 are false positive results for the automated analyzer. The number below each bar is the number of samples represented by the bar.

### VRC Cohort

#### Samples and Demographics

To further examine automated HSV serology assay performance, we took advantage of a unique cohort of 1017 persons with genital HSV infection confirmed by PCR or culture through genital herpes studies at the UW VRC from 1981 to 2019. For this study, we included samples from persons with genital herpes with PCR- or culture-confirmed HSV infection who had serological testing using the WB at least 6 months (median 5.8 years, range 6 months to 50 years) after the initial episode of genital herpes. Given the focus on genital herpes, 886 (87%) people in this cohort had PCR- or culture-confirmed HSV-2 infections and 131 (13%) people had PCR- or culture-confirmed genital HSV-1 infections. WB results were 100% concordant with the PCR or culture results. Of the 886 people with confirmed HSV-2 genital infections, 320 (36%) also had HSV-1 antibodies detected on the western blot, most likely from prior HSV-1 infection.

The VRC cohort was comprised of 669 women (65.8%) and 348 men (34.2%) who provided samples between 1981 and 2019 (median 1989). Most people were white (86.3%), and the overall median age was 34.3 (range: 16-88 years, Table 4 and Figure S2).

**Table 4:**
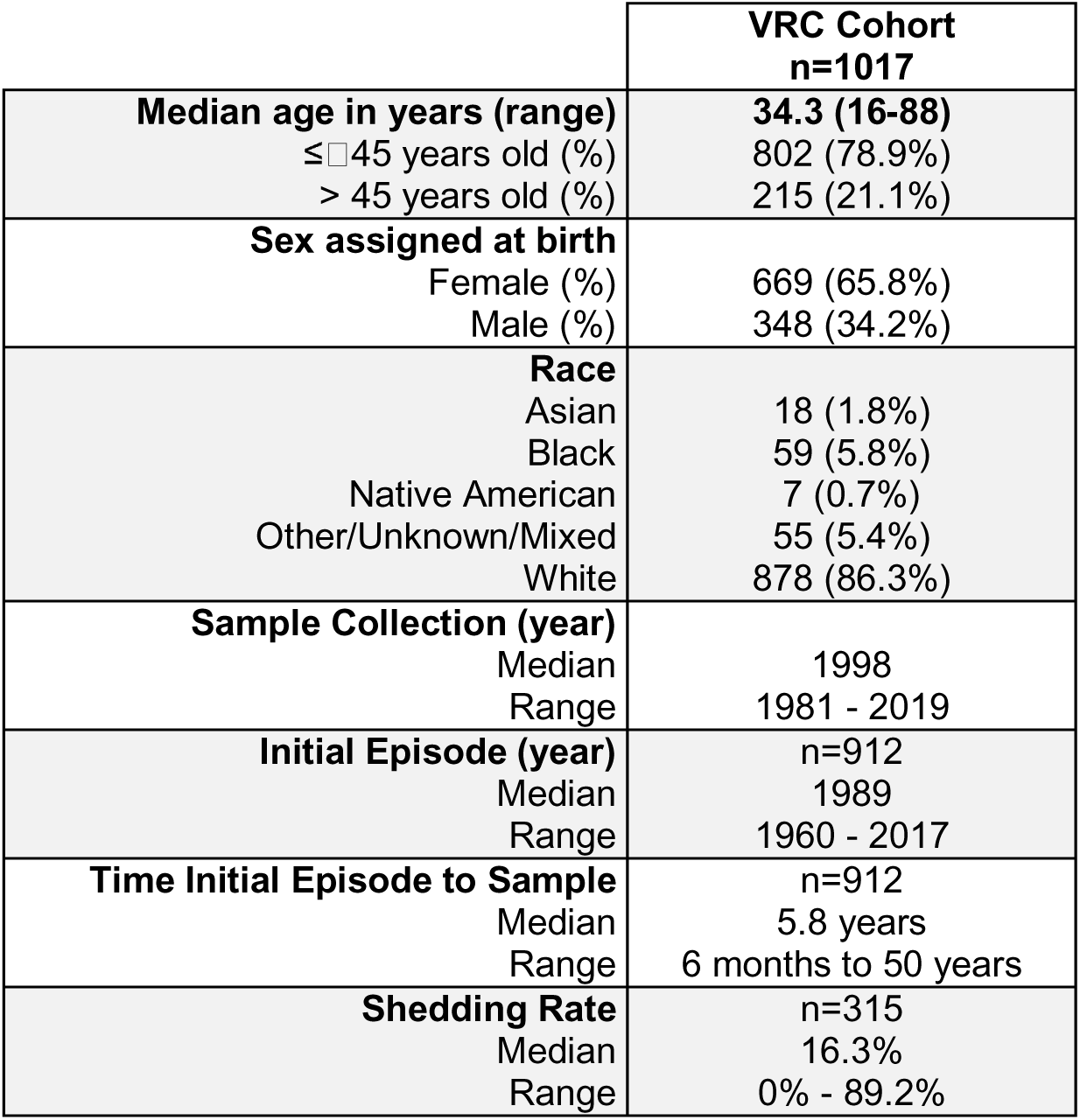
Demographic information for the VRC cohort. Time since initial episode and shedding rate information are only available for a subset of persons in this cohort.

|  | VRC Cohort<br>n=1017 |
| --- | --- |
| **Median age in years (range)** | **34.3 (16-88)** |
| ≤ 45 years old (%) | 802 (78.9%) |
| > 45 years old (%) | 215 (21.1%) |
| **Sex assigned at birth** |  |
| Female (%) | 669 (65.8%) |
| Male (%) | 348 (34.2%) |
| **Race** |  |
| Asian | 18 (1.8%) |
| Black | 59 (5.8%) |
| Native American | 7 (0.7%) |
| Other/Unknown/Mixed | 55 (5.4%) |
| White | 878 (86.3%) |
| **Sample Collection (year)** |  |
| Median | 1998 |
| Range | 1981 - 2019 |
| **Initial Episode (year)** | n=912 |
| Median | 1989 |
| Range | 1960 - 2017 |
| **Time Initial Episode to Sample** | n=912 |
| Median | 5.8 years |
| Range | 6 months to 50 years |
| **Shedding Rate** | n=315 |
| Median | 16.3% |
| Range | 0% - 89.2% |

#### Assay Performance

The VRC cohort is comprised of samples that are positive for HSV-1 or HSV-2 antibodies based both on WB results and viral confirmation of genital infection. In this cohort, all automated instruments had lower sensitivity and specificity for detecting HSV-1 IgG with all instruments having a sensitivity <90%. Conversely, in this population, the automated assays are more sensitive for detecting HSV-2 antibodies than in a typical screening population (Table 5). However, the automated assays had lower specificity for HSV-2 IgG in this cohort. When only analyzing samples with PCR- or culture-confirmed HSV-1 or HSV-2 genital infection (e.g. only positive for either HSV-1 IgG (131 samples) *or* HSV-2 IgG (566 samples)), the sensitivities of the HSV-1 IgG assays were not improved (80.6% for Bio-Rad, 87.8% for DiaSorin, and 69.5% for Roche). For HSV-2, all instruments’ sensitivities were largely unchanged and were within 0.6% of their values from the full cohort Assay specificities for this subset were unchanged compared to the full cohort (Table S1).

**Table 5:**
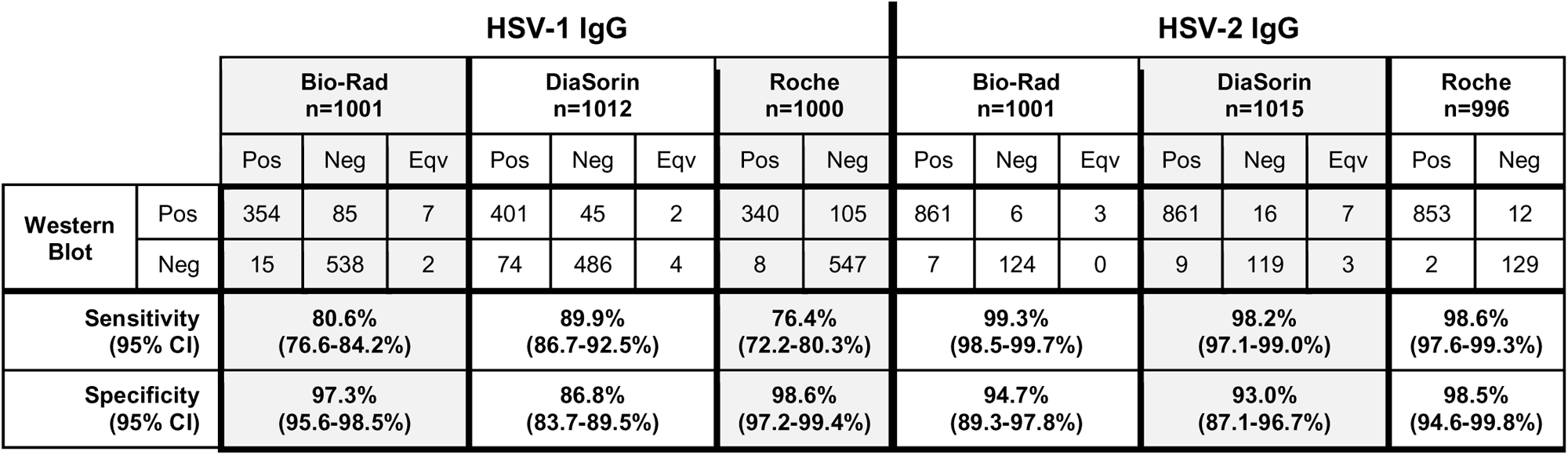
Test results for each automated instrument compared to the western blot results for the VRC cohort. Equivocal (Eqv) results were not included in calculating sensitivity and specificity.

Overall, there were 16 (13 for HSV-1 IgG, 3 for HSV-2 IgG) equivocal results on the Bio-Rad instrument and 34 (18 for HSV-1 IgG, 16 for HSV-2 IgG) on the DiaSorin instrument. In contrast to the clinical testing cohort where most of the Bio-Rad results had non-equivocal final results, only 25% of the Bio-Rad equivocal results in the VRC cohort had a non-equivocal result after repeating. All 4 revised results became positive for HSV-1 IgG, yielding a false positive result in 2 cases. For DiaSorin, 53% (12 HSV-1 IgG, 6 HSV-2 IgG) of the equivocal results from this cohort had a non-equivocal final result with 11 results concordant with the WB and the remaining 7 results falsely positive for HSV-1 IgG. All equivocal samples were unique and no samples from the VRC cohort were equivocal for multiple assays or instruments.

Patterns of discordant results for the VRC cohort were similar to those for the clinical testing cohort (Figure 4). Overall, the DiaSorin instrument alone accounted for at least half of the total false positives for either HSV-1 IgG (78%) or HSV-2 IgG (50%). There was a trend for the VRC cohort to have a larger proportion of samples test falsely positive (13.3% for HSV-1 IgG, 10.7% for HSV-2 IgG) compared to the clinical testing cohort (10.2% for HSV-1 IgG and 6.5% for HSV-2 IgG), likely reflecting the lack of double negative samples in this cohort. There were significantly more HSV-1 IgG false negative results in the VRC cohort than the clinical testing sample remnant cohort (25.9% vs. 16.7%, p-value=0.002). Conversely, there were significantly fewer HSV-2 false negative results in the VRC cohort than the clinical testing cohort (3.1% vs. 16.4%, p-value <0.001).

**Figure 4:**
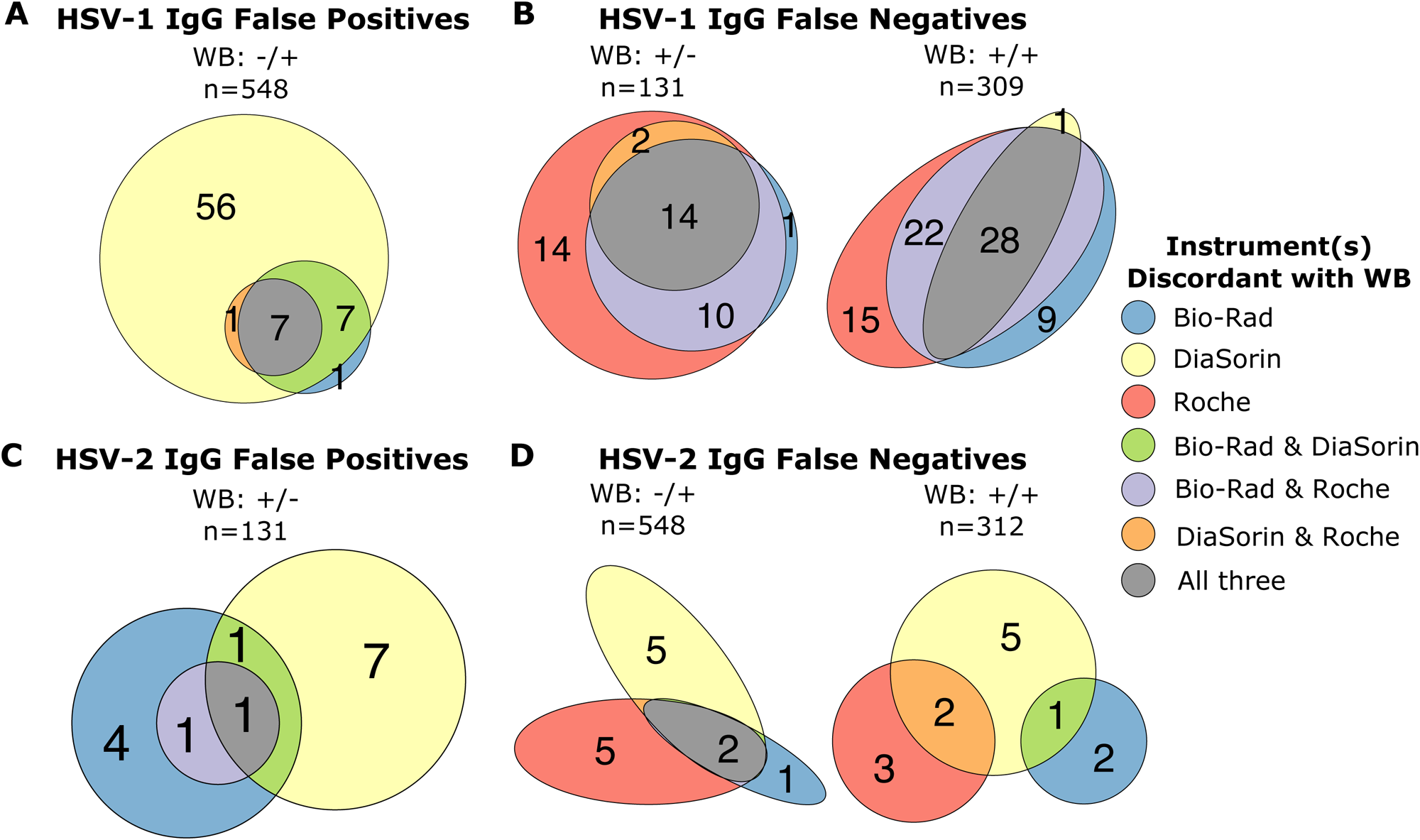
Euler plots showing the number of samples from the VRC cohort that disagree with the western blot results for each automated instrument colored by which instruments were discordant with the WB results. Data are presented as in Figure 1. A) Number of samples that were negative for HSV-1 IgG by WB, but falsely positive on the automated instruments. B) Number of samples that were positive for HSV-1 IgG by WB, but falsely negative on the automated instruments. C) Number of samples that were negative for HSV-2 IgG by WB, but falsely positive on the automated instruments. D) Number of samples that were positive for HSV-2 IgG by WB, but falsely negative on the automated instruments. Plots made using the eulerr R package.^22^

#### Antibody indices compared to days since initial episode

All serum samples in the VRC cohort were collected at least 6 months after a person’s initial episode of genital herpes (range 6 months to 50 years). There was a small but statistically significant positive correlation between the number days after initial episode of genital herpes that a sample was collected and the quantitative index value for both HSV-1 IgG and HSV-2 IgG on all instruments (Table 6).

**Table 6:**
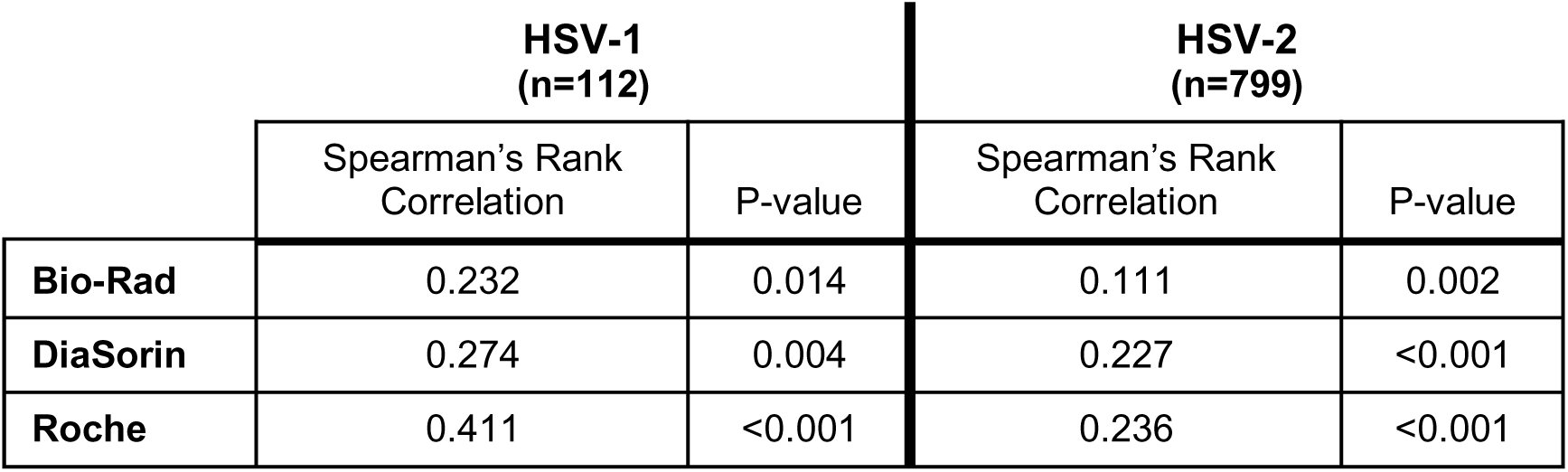
Spearman’s rank correlation for HSV-1 IgG or HSV-2 IgG quantitative index and days since initial genital episode. This analysis corrects for the effects of both age and collection date.

### Estimates of Test Characteristics

We calculated the sensitivities and specificities from all 1996 samples to estimate the overall performance of these assays (Table 7). Using the 2015-2016 NHANES population prevalence in the United States for persons aged 14-49 for HSV-1 of 48% and HSV-2 of 12%,^3^ we calculated the positive and negative predictive value for each assay based on the sensitivities and specificities from all samples tested (Table 7) and modeled the effect of changes in prevalence on the PPV and NPV (Figure 5).

**Figure 5:**
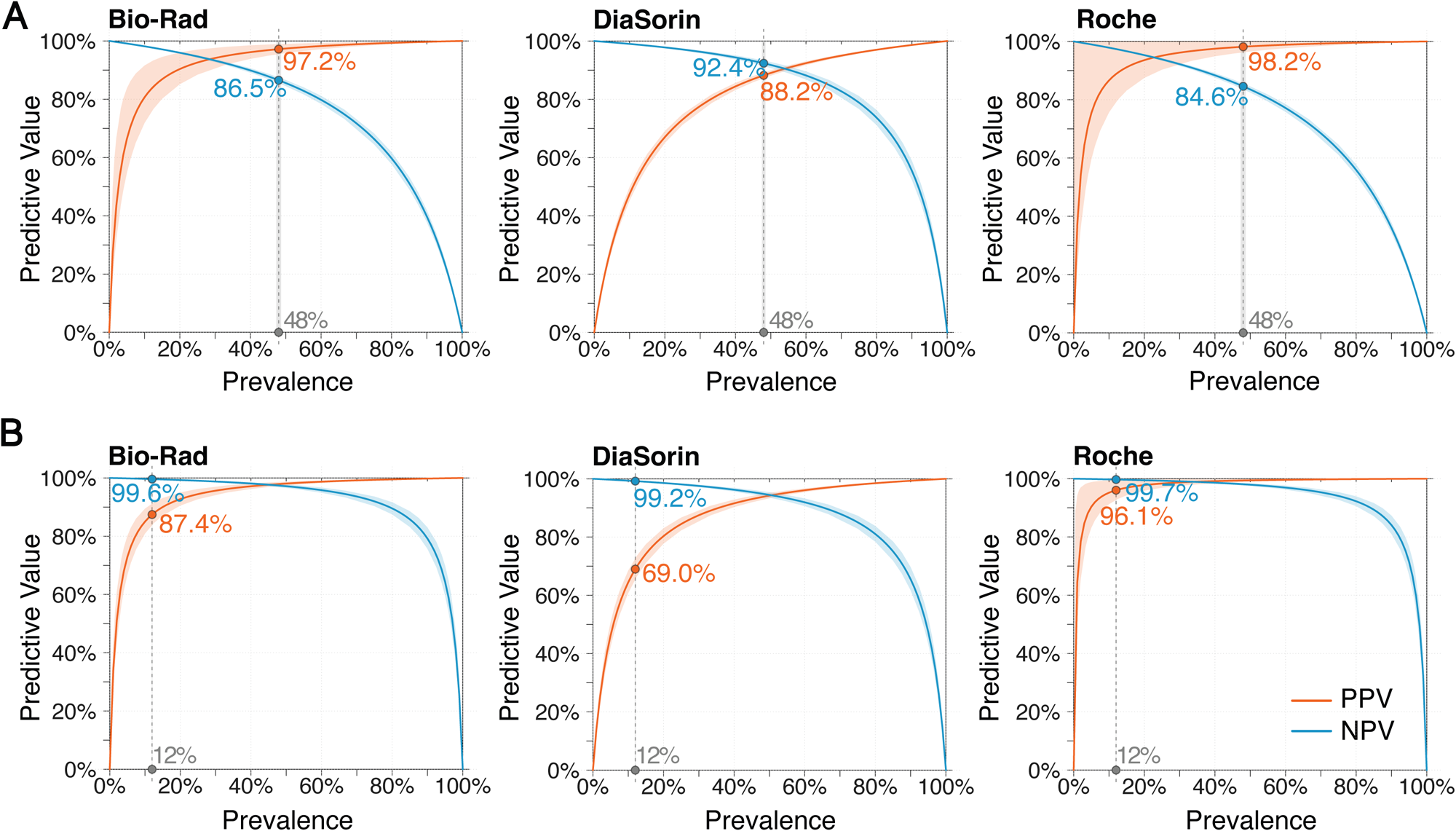
The positive and negative predictive values for each assay as a function of prevalence based on the sensitivities and specificities calculated from all samples tested. A) Modeling of the positive and negative predictive values by prevalence for each HSV-1 IgG assay. The grey line indicates a population prevalence for HSV-1 in the US of 48%. ^3^ B) Modeling of the positive and negative predictive values by prevalence for each HSV-2 IgG assay. The grey line indicates the expected population prevalence for HSV-2 in the US of 12%. For all plots, the shaded areas represent the percent uncertainty for the sensitivity and specificity, set to the 95% CI of either the sensitivity or specificity, whichever was larger.

**Table 7:** Sensitivity (SN) and specificity (SP) for all instruments and all cohorts, including all samples combined.

|  | HSV-1 IgG |  |  |  |  |  | HSV-2 IgG |  |  |  |  |  |
| --- | --- | --- | --- | --- | --- | --- | --- | --- | --- | --- | --- | --- |
|  | Bio-Rad |  | DiaSorin |  | Roche |  | Bio-Rad |  | DiaSorin |  | Roche |  |
|  | SN | SP | SN | SP | SN | SP | SN | SP | SN | SP | SN | SP |
| All Samples<br>n=1996 | 83.5% | 97.8% | 92.1% | 88.7% | 80.5% | 98.6% | 97.1% | 98.1% | 94.5% | 94.2% | 97.9% | 99.5% |
| Clinical<br>Testing<br>Samples<br>n=979 | 87.1% | 98.2% | 94.8% | 90.4% | 85.9% | 98.7% | 91.9% | 98.8% | 85.4% | 94.5% | 96.1% | 99.7% |
| VRC Samples<br>n=1017 | 80.6% | 97.3% | 89.9% | 86.8% | 76.4% | 98.6% | 99.3% | 94.7% | 98.2% | 93.0% | 98.6% | 98.5% |

Due to the higher prevalence of HSV-1, the positive predictive values are higher for all HSV-1 IgG assays despite those assays having worse sensitivity and specificity than the HSV-2 IgG assays. Due to the low prevalence of HSV-2, ∼1 in 26 positive HSV-2 results on the Roche assay are false positives, while the likelihood that a positive HSV-2 result is a false positive are even higher for the other assays: ∼1 in 8 for Bio-Rad and nearly 1 in 3 for DiaSorin.

Using the quantitative data, we also looked at if a different quantitative index threshold may be better for maximizing each assay’s sensitivity and specificity. Based on maximizing the receiver operating characteristic curves (Figure S3), for the DiaSorin assays increasing the threshold to 1.725 for HSV-1 IgG or 1.135 for HSV-2 IgG would give the optimal specificity and sensitivity. For both the Bio-Rad and Roche instruments, decreasing the threshold would yield the optimal specificity and sensitivity for the HSV-1 and HSV-2 IgG assays. However, decreasing the threshold would result in a lower specificity and more false positive results.

## DISCUSSION

In this large-scale comparison of HSV serologic diagnostic methods, we found substantial differences between the performance of three commonly used, automated FDA-cleared immunoassays for detecting HSV-1 and HSV-2 IgG manufactured by Bio-Rad, DiaSorin, and Roche. For HSV-1 IgG, there was a clear trade-off between sensitivity and specificity for all assays. The DiaSorin HSV-1 IgG assay had the highest sensitivity, but lowest specificity. On the other hand, the Bio-Rad and Roche HSV-1 IgG assays had poor sensitivities (<85%), but higher specificities (>97%). For HSV-2 there was a clear best assay with the Roche HSV-2 IgG assay having both the highest sensitivity (97.9%) and highest specificity (99.5%). The Bio-Rad HSV-2 IgG assay was nearly as good as the Roche assay, but the DiaSorin HSV-2 assay performed worst for both sensitivity and specificity. Unfortunately for laboratories, there does not seem to be a one-size-fits-all approach to automated HSV serology assays. Indeed, while our study indicates that at least some automated HSV-2 IgG assays have promising performance metrics, none of the HSV-1 IgG assays tested are adequate.

A notable aspect of our results is that we found higher specificities for HSV-2 than previous studies.^14^ Most prior studies compared results to ELISAs, most commonly the HerpeSelect test by Focus Diagnostics (purchased by DiaSorin in 2016). To our knowledge, this is the first large-scale comparison of a Roche HSV-1 IgG assay and first comparison of any Roche HSV-2 IgG assay since 1999.^17^ Similarly, the last large-scale comparisons of the Bio-Rad HSV-1 and HSV-2 IgG assays to other HSV serologic diagnostic methods were published in 2010-11.^18,19^

A strength of this study includes comparing automated HSV serology assays using samples not only measured by HSV WB, but also with virologic confirmation of HSV infection. The VRC cohort provides a set of samples with a high degree of confidence in their diagnosis that serve as an excellent cohort for determining the optimal performance characteristics for these assays. Indeed, we found that the automated assays all performed quite well at detecting HSV-2 antibodies in these samples, albeit with the DiaSorin assay still having the worst performance characteristics. However, the automated assays struggled to detect HSV-1 IgG. While it is encouraging to find assays that perform better than expected at detecting HSV-2 antibodies, as HSV-1 continues to increase in prevalence as a cause of genital herpes, improved serologic diagnostics are needed for both HSV-2 and HSV-1.

In December 2023, DiaSorin notified customers that current HSV-2 reagent lots, including those used in this study, may have increased numbers of equivocal or false positive results. In our study, compared to the Diasorin HSV-1 assay, the DiaSorin HSV-2 assay did not seem to be enriched for equivocal or false-positive results, as the HSV-2 assay had fewer equivocal (16) or false positive (43) final results than the HSV-1 assay (21 equivocal and 133 false positive results). Of note, our data also confirms the high false positive rate of the DiaSorin HSV-2 assay seen in Jan-Feb 2021 by Quest, especially for specimens with low-positive index values.^20^ Given the performance issues of the DiaSorin assay predate the manufacture of the lots subject to the notification, further consideration for the utility of the assay is warranted. Based on our ROC analyses, even if the threshold for the DiaSorin assays were increased, the best sensitivity and specificity for their HSV-2 assay would remain worse than the other assays tested.

Limitations of this study include the use of clinical remnants rather than fresh prospectively collected specimens. Most (86%) of the VRC cohort specimens were stored frozen for >10 years, though sensitivity analysis of testing performance by date of collection demonstrated no substantial difference in assay sensitivity for samples collected prior to 2000 (n=555) vs after 2000 (n=462) (Table S2). This study was not specifically designed to assess the effects of demographics on assay performance, but, notably, we found mostly minor differencs in the performance of the different assays by patient age in the clinical remnant cohort (Table S3, Supplemental Material). The VRC cohort was less diverse than the clinical remnant cohort and only included HIV-negative individuals with genital herpes, which may limit generalizability to other testing contexts. Our study also included 16 samples from persons under 18 years old, although none of the automated assays are FDA-cleared for pediatric testing. Specimens were tested on the Bio-Rad instrument after one additional freeze-thaw compared to DiaSorin and Roche and 47 of the equivocal samples were also repeated after an additional freeze-thaw, which may have slightly impacted test performance. Though the Roche assay demonstrated perfect specificity in our WB double negative cohort, only ∼430 specimens were tested in this set.

Taken together, our study demonstrates that currently available HSV serologic diagnostics have limitations precluding widespread use for HSV-1 and HSV-2 testing.^14,21^ In our hands, the Roche HSV-2 assay performed the best, and, overall, the Roche HSV assays would minimize potential harm from false positives as neither assay had any false positives among individuals seronegative for both HSV subtypes by WB. In addition, the Roche assay does not have equivocal results and performs well across the index value range, as opposed to the DiaSorin and Bio-Rad assays which have lower specificity at low index values and for which additional testing is required to resolve equivocal results. Overall, further work is required to adjudicate type-specific accuracy and reduce cross-reactivity among automated assays. Given the role that adsorption plays in resolving indeterminate results on the WB, it is notable that no adsorption reagents are available for automated HSV serology assays. Future work should examine how adsorption reagents could similarly resolve low positive index values on automated assays. Further improvements and investments in HSV serologic diagnostics remain necessary to support both clinical care and potential future screening.

## Supporting information

Supplemental Data - Line-item Testing

Supplemental Data 2 - Linearity

## Data Availability

All data produced in the present work are contained in Supplemental Data.

## Acknowledgements

We thank the staff at the University of Washington Clinical Virology, Retrovirology, and Immunology labs for their support of this project, especially Grace Matias, Audrey Ruhland, and the staff and participants at the University of Washington Virology Research Clinic.

## References

1. Wald, A. et al. Reactivation of genital herpes simplex virus type 2 infection in asymptomatic seropositive persons. N. Engl. J. Med. 342, 844–850 (2000).

2. James, C. et al. Herpes simplex virus: global infection prevalence and incidence estimates, 2016. Bull. World Health Organ. 98, 315–329 (2020).

3. McQuillan, G., Kruszon-Moran, D., Flagg, E. W. & Paulose-Ram, R. Prevalence of Herpes Simplex Virus Type 1 and Type 2 in Persons Aged 14-49: United States, 2015-2016. NCHS Data Brief 1–8 (2018).

4. Ryder, N., Jin, F., McNulty, A. M., Grulich, A. E. & Donovan, B. Increasing role of herpes simplex virus type 1 in first-episode anogenital herpes in heterosexual women and younger men who have sex with men, 1992-2006. Sex. Transm. Infect. 85, 416–419 (2009).

5. Nath, P., Kabir, M. A., Doust, S. K. & Ray, A. Diagnosis of Herpes Simplex Virus: Laboratory and Point-of-Care Techniques. Infect. Dis. Rep. 13, 518–539 (2021).

6. Koelle, D. M., Benedetti, J., Langenberg, A. & Corey, L. Asymptomatic reactivation of herpes simplex virus in women after the first episode of genital herpes. Ann. Intern. Med. 116, 433–437 (1992).

7. Johnston, C. Diagnosis and Management of Genital Herpes: Key Questions and Review of the Evidence for the 2021 Centers for Disease Control and Prevention Sexually Transmitted Infections Treatment Guidelines. Clin. Infect. Dis. Off. Publ. Infect. Dis. Soc. Am. 74, S134–S143 (2022).

8. McGeoch, D. J., Moss, H. W., McNab, D. & Frame, M. C. DNA sequence and genetic content of the HindIII l region in the short unique component of the herpes simplex virus type 2 genome: identification of the gene encoding glycoprotein G, and evolutionary comparisons. J. Gen. Virol. 68 (Pt 1), 19–38 (1987).

9. US Preventive Services Task Force et al. Serologic Screening for Genital Herpes Infection: US Preventive Services Task Force Recommendation Statement. JAMA 316, 2525–2530 (2016).

10. Workowski, K. A. et al. Sexually Transmitted Infections Treatment Guidelines, 2021. MMWR Recomm. Rep. Morb. Mortal. Wkly. Rep. Recomm. Rep. 70, 1–187 (2021).

11. Ashley, R. L., Militoni, J., Lee, F., Nahmias, A. & Corey, L. Comparison of Western blot (immunoblot) and glycoprotein G-specific immunodot enzyme assay for detecting antibodies to herpes simplex virus types 1 and 2 in human sera. J. Clin. Microbiol. 26, 662–667 (1988).

12. Ashley, R. L. & Wald, A. Genital herpes: review of the epidemic and potential use of type-specific serology. Clin. Microbiol. Rev. 12, 1–8 (1999).

13. Wald, A. & Ashley-Morrow, R. Serological testing for herpes simplex virus (HSV)-1 and HSV-2 infection. Clin. Infect. Dis. Off. Publ. Infect. Dis. Soc. Am. 35, S173–182 (2002).

14. Agyemang, E. et al. Performance of Commercial Enzyme-Linked Immunoassays for Diagnosis of Herpes Simplex Virus-1 and Herpes Simplex Virus-2 Infection in a Clinical Setting. Sex. Transm. Dis. 44, 763–767 (2017).

15. Tronstein, E. et al. Genital shedding of herpes simplex virus among symptomatic and asymptomatic persons with HSV-2 infection. JAMA 305, 1441–1449 (2011).

16. RStudio Team. RStudio: Integrated Development Environment for R. PBC (2022).

17. Groen, J. et al. Evaluation of a fully automated glycoprotein G-2 based assay for the detection of HSV-2 specific IgG antibodies in serum and plasma. J. Clin. Virol. Off. Publ. Pan Am. Soc. Clin. Virol. 12, 193–200 (1999).

18. Binnicker, M. J., Jespersen, D. J. & Harring, J. A. Evaluation of three multiplex flow immunoassays compared to an enzyme immunoassay for the detection and differentiation of IgG class antibodies to herpes simplex virus types 1 and 2. Clin. Vaccine Immunol. CVI 17, 253–257 (2010).

19. LeGoff, J. et al. Performance of the BioPlex 2200 multiplexing immunoassay platform for the detection of herpes simplex virus type 2 specific antibodies in African settings. Clin. Vaccine Immunol. CVI 18, 1191–1193 (2011).

20. Prince, H. E., Batterman, H. J. & Marlowe, E. M. Characterization of Serum Samples With Discordant Results in 2 Herpes Simplex Virus Type 2 IgG Assays. Sex. Transm. Dis. 49, 353–359 (2022).

21. Mujugira, A. et al. Performance of the Focus HerpeSelect-2 enzyme immunoassay for the detection of herpes simplex virus type 2 antibodies in seven African countries. Sex. Transm. Infect. 87, 238–241 (2011).

22. Larsson, J. eulerr: Area-Proportional Euler and Venn Diagrams with Ellipses. (2022).

