## Supplemental Figure 1 for "Performance characteristics of highly automated HSV-1 and HSV-2 IgG testing"

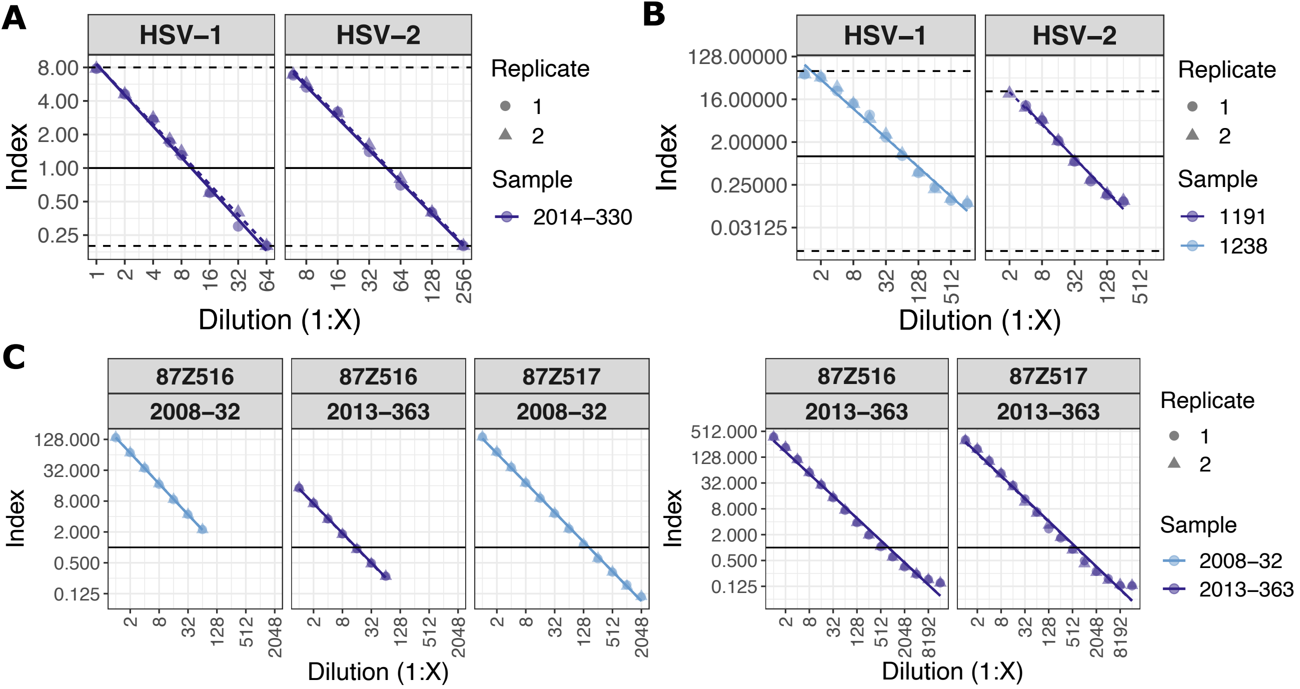


**Figure S1:** Dilutions of high-positive samples show the typical linearity range for each assay. The solid horizontal line indicates an index of 1.0. Dashed lines denote the limits of an instrument's reportable range, where applicable. A) Linearity for the Bio-Rad HSV-1 and HSV-2 IgG assays using the same sample (2014-330) for both HSV-1 and HSV-2. B) Linearity studies for the DiaSorin HSV-1 and HSV-2 IgG assays. Different samples were used for HSV-1 and HSV-2. C) Linearity studies for both instruments that ran the Roche HSV-1 and HSV-2 IgG assays. The Roche assays were the only ones run on multiple instruments. For instrument 87Z516 HSV-1 IgG testing, two samples were run to better cover the lower end of the index range.
