## Supplemental Figure 2 for "Performance characteristics of highly automated HSV-1 and HSV-2 IgG testing"

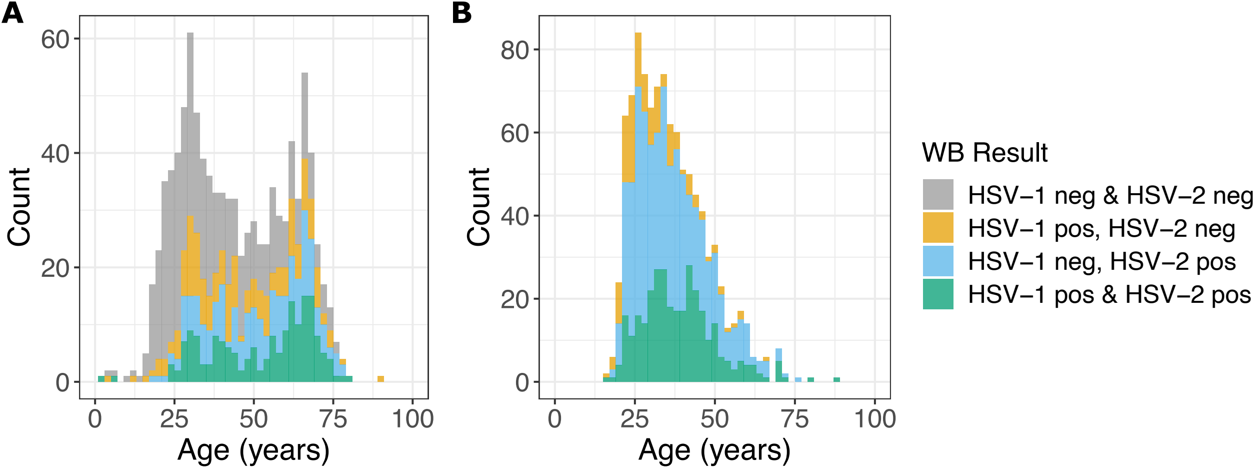


**Figure S2:** The age distributions for the sample cohorts. A) The clinical testing sample remnant cohort clearly shows a bimodal age distribution. As expected for serologic testing for a life-long viral infection, the number of persons positive for HSV-1 and/or HSV-2 antibodies also increases with age. B) The VRC cohort age distribution is unimodal and only the younger population is present.
