## Supplemental Figure 3 for "Performance characteristics of highly automated HSV-1 and HSV-2 IgG testing"

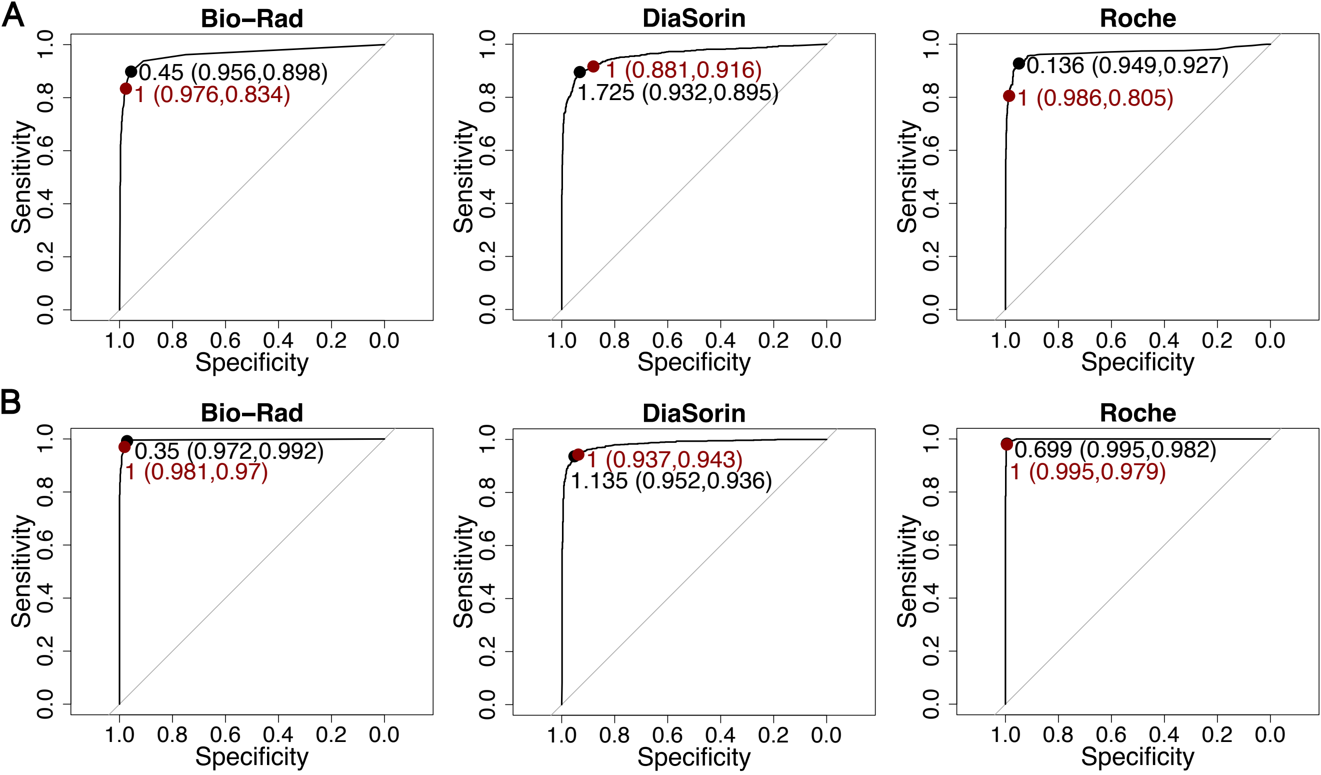


**Figure S3:** Receiver operator characteristic (ROC) curves for all samples and all instruments. A) ROC curves for the HSV-1 IgG assays. B) ROC curves for the HSV-2 IgG assays. The red point shows the threshold of 1.0 used in this study and the resulting specificity, and sensitivity (in that order) when not excluding equivocal results. For the Bio-Rad and DiaSorin instruments, these values differ slightly from those presented in the main text (Table 7) where equivocal results were excluded from calculations of sensitivity and specificity. The black point shows the optimal threshold for each assay as calculated using Youden's J statistic. The R pROC package was used to construct these plots [Sup. Ref. 3].
