## Supplemental Material for "Performance characteristics of highly automated HSV-1 and HSV-2 IgG testing"

*Equivocal samples in clinical testing remnant sample cohort*

For the 19 samples that initially resulted as equivocal on the Bio-Rad instrument, 8 were equivocal for HSV-1 IgG and 11 for HSV-2 IgG. The majority of these equivocal results—7 of 8 for HSV-1 IgG and 7 of 11 for HSV-2 IgG—occurred when a sample was positive for the tested antibodies by WB. Interestingly, none of the Bio-Rad instrument’s equivocal results were for samples seronegative for both HSV-1 and HSV-2 antibodies by WB. Bio-Rad recommends collecting new specimens for repeat testing for patients with equivocal results. We could not do that for this study and instead repeated all equivocal results at least twice. The median result was accepted as the final result. For this clinical testing remnant sample cohort, the majority of equivocal results (7 of 8 for HSV-1 IgG and 7 of 11 for HSV-2 IgG) were revised to a non-equivocal final result after repeat testing. 8 of these samples (5 for HSV-1 and 3 for HSV-2) were correctly revised to positive results while the remaining 6 revised results were corrected to results discordant with the WB with 3 false positive HSV-2 IgG results, 1 false positive HSV-1 IgG result, 1 false negative HSV-1 result, and 1 false negative HSV-2 result.

For the DiaSorin assay, all 24 initially equivocal results—16 for HSV-1 and 8 for HSV-2—were repeated in duplicate. Samples yielding equivocal results on the DiaSorin instrument came from all WB categories (9 were HSV-1 IgG and HSV-2 IgG seronegative by WB, 4 were HSV-1 IgG positive only, 7 were HSV-2 IgG positive only, and 4 were HSV-1 IgG and HSV-2 IgG seropositive). Overall, the DiaSorin instrument tended to report equivocal results for samples that were negative for the tested antibodies by WB (12 of 16 samples for HSV-1 and 5 of 8 for HSV-2). Again, the median result was accepted as the final result. Of these initially equivocal results, only 3 samples—1 for HSV-1 and 2 for HSV-2—yielded non-equivocal final results and all 3 samples were correctly revised to negative results.

*Equivocal samples in the VRC cohort*

With the VRC cohort, the Bio-Rad instrument initially had 16 equivocal results—13 HSV-1 IgG and 3 HSV-2 IgG. The equivocal HSV-1 IgG results came from all WB result categories in this cohort (2 HSV-1 IgG positive only, 4 HSV-2 IgG positive only, and 7 positive for HSV-1 and HSV-2 IgG by WB) and the 3 equivocal HSV-2 results were only from samples positive for HSV-2 IgG by WB (2 positive for HSV-2 IgG only and 1 positive for HSV-1 and HSV-2 IgG). Four of the equivocal HSV-1 IgG results were revised to positive after repeating. For 2 of these samples, this was a false positive result. The remaining 12 equivocal results from the Bio-Rad instrument remained equivocal after repeating. The DiaSorin instrument had 34 equivocal results. All 18 samples equivocal for HSV-1 IgG were positive for HSV-2 IgG by WB (14 samples were positive for HSV-2 IgG only and 4 were positive for HSV-1 and HSV-2 IgG). The 16 samples with HSV-2 IgG equivocal results were from all 3 WB categories represented in this cohort (3 samples were HSV-1 IgG positive only, 7 were HSV-2 IgG positive only, and 6 were HSV-1 and HSV-2 IgG positive by WB). Eighteen results—12 for HSV-1 IgG and 6 for HSV-2 IgG—were revised to a non-equivocal result. Only 5 of the revised HSV-1 IgG results were concordant with the WB and the remaining 7 yielded false positive HSV-1 IgG results. All 6 of the revised HSV-2 IgG results were finalized to positive results in concordance with the WB.

*Effects of demographics on assay performance for the clinical testing sample remnant cohort*

We also examined if any of the automated assays performed better on any subpopulation defined by our study demographics (Table S3). Consistent with prior studies, older individuals had higher seroprevalence by WB for both HSV-1 and, especially, HSV-2 compared to people ≤45 years old.^1^ Specimens from women or from Hispanic or non-white individuals also had higher positivity for HSV-1 or HSV-2 compared to men and white, non-Hispanic individuals.^1,2^ Although test sensitivity and specificity should be unaffected by disease prevalence, for both women and Hispanic or non-white individuals, the increase in population prevalence coincided with equivalent or higher sensitivity, but equivalent or slightly lower specificity when compared to men or white, non-Hispanic individuals, respectively. This pattern did not hold for the older age group, which had lower sensitivities and specificities for all HSV-1 assays. Both age groups had generally equivalent specificity for all HSV-2 assays, but the Bio-Rad HSV-2 assay had a lower sensitivity for the older age group and the DiaSorin and Roche HSV-2 assays had a higher sensitivity for individuals >45 years old.

**References**

1.	Bradley, H., Markowitz, L. E., Gibson, T. & McQuillan, G. M. Seroprevalence of herpes simplex virus types 1 and 2--United States, 1999-2010. *J. Infect. Dis.* 209, 325–333 (2014).

2.	Smith, J. S. & Robinson, N. J. Age-specific prevalence of infection with herpes simplex virus types 2 and 1: a global review. *J. Infect. Dis.* 186 Suppl 1, S3-28 (2002).

3.	Robin, X. *et al.* pROC: an open-source package for R and S+ to analyze and compare ROC curves. *BMC Bioinformatics* 12, 77 (2011).
